# Risk of death by age and gender from CoVID-19 in Peru, March-May, 2020

**DOI:** 10.1101/2020.06.14.20123315

**Authors:** Cesar Munayco, Gerardo Chowell, Amna Tariq, Eduardo A. Undurraga, Kenji Mizumoto

## Abstract

Peru implemented strict social distancing measures during the early phase of the epidemic and is now experiencing one of the largest CoVID-19 epidemics in Latin America. Estimates of disease severity are an essential indicator to inform policy decisions about the intensity and duration of interventions needed to mitigate the outbreak. Here we derive delay-adjusted case fatality rates (aCFR) of CoVID-19 in a middle-income country in South America.

We used government-reported time series of CoVID-19 cases and deaths stratified by age group and gender.

Our estimates as of May 25, 2020, of the aCFR for men and women are 10.8% (95%CrI: 10.5–11.1%) and 6.5% (95%CrI: 6.2–6.8%), respectively, and an overall aCFR of 9.1% (95%CrI: 8.9–9.3%). Our results show that senior individuals are the most severely affected by CoVID-19, particularly men, with aCFR of almost 60% for those aged 80-years. We found that men have a significantly higher cumulative morbidity ratio than women across most age groups (proportion test, p-value< 0.001), with the exception of those aged 0–9 years.

The COVID-19 epidemic is imposing a large mortality burden in Peru. Senior individuals, especially those who are older than 70 years of age, are being disproportionately affected by the COVID-19 pandemic.

## Introduction

As of May 25, 2020, more than 5.5 million CoVID-19 (illness caused by the novel coronavirus SARS-COV-2) cases and ∼340,000 deaths have been reported from almost every country and territory around the globe [1, 2]. The CoVID-19 pandemic has imposed a substantial burden on health systems, economies, and societies globally, and there are strong indicators pointing to a disproportionate impact on low- and middle-income countries [3–5]. Since its initial outbreak in China, the world has tracked the CoVID-19 pandemic proliferating across Europe and Asia, seeding hotspots in North America, and the Middle East [6]. The pandemic arrived late to South America; Brazil reported its first case on February 26, 2020 [7]. Neighboring countries contracted CoVID-19 cases in subsequent days; South America has registered more than 600,000 cases and 30,600 deaths as of May 24, 2020 [1]. Although many South American countries imposed strict control measures, including travel bans, school closures, and lockdowns early in the epidemic, the pandemic now rivals Europe in size, with COVID-19 cases and death counts increasing rapidly [1, 5]. Other factors, including high poverty rates, informal economies, inadequate water, sanitation, and hygiene infrastructure, frail healthcare systems, and insufficient medical supplies, further complicate the health and socioeconomic effects of the CoVID-19 pandemic [5, 8–10]. Governments in South America are now facing the social and economic consequences of SARS-COV-2 containment measures, while struggling to the contain the virus [9].

Peru, a country of about 30 million people, is experiencing one of the largest CoVID-19 epidemics in Latin America. With an approximate two-fold increase in the cumulative case count during the past two weeks, Peru has reported almost 129,148 cases and 7660 deaths as of May 25, 2020 [11]. The majority (63%) of COVID-19 cases have been confirmed in Lima, the capital of Peru [11]. The government of Peru initiated social distancing measures soon after the confirmation of the first imported case in Peru on March 6, 2020 [12]. The initial epidemic control measures included school closures on March 11, 2020 followed by the suspension of large gatherings and flights from Europe and Asia the next day. Subsequently the government declared national emergency in the country and closed country borders on March 16, 2020 [13]. Despite these forthcoming and swift control measures, community transmission was reported to be occurring by March 17, 2020, forcing the implementation of a night time curfew as of March 18, 2020 [13].

Estimates from the early stage of the epidemic in Peru (March 2020) showed sustained transmission in Lima with an estimated of reproduction number R∼2.3 (95% CI: 2.0, 2.5) [14]. Moreover, the 20-days ahead forecast for Lima indicated that the prompt social distancing measures have significantly slowed down the initial spread of the virus in the region [14]. The crude case fatality rate (CFR), defined as the number of cumulative deaths and cases at a single time point, in Peru is estimated at 5.9%, approximating the global crude CFR average of ∼6.3% [15]. Statistical analyses and mathematical models using data from Peru suggest that under current epidemic growth, the number of CoVID-19 infected individuals could surpass the healthcare system capacity [16].

The clinical spectrum of CoVID-19 ranges from asymptomatic cases to clinical conditions characterized by respiratory failure, to multiorgan and systemic manifestations which can cause death [17–19]. CFR is a statistical entity used to estimate severity of the epidemic [20] as it provides a criterion for the public health officials to gauge the intensity and duration of public health interventions required to contain the epidemic [21]. However, it becomes challenging to estimate CFR during an epidemic as CFR estimates are sensitive to right censoring of the data that occurs because of the time lag between the date of symptoms onset and the date of death for a case [22–24]. Moreover, under-reporting of cases because mild or asymptomatic cases can go undetected by disease surveillance systems also underestimates CFR [22, 25].

In this study we provide real-time estimates of adjusted age-specific CFR during the CoVID-19 epidemic in Peru, through May 25, 2020 to assess the severity of the SARS-CoV-2 epidemic in the region. The country has been performing PCR tests with the average positivity rate estimated at ∼8.6% as of March 30, 2020 [14]. In Peru ∼85% of ICU beds with ventilators are currently occupied by patients [26], therefore our current estimates are not affected by excess deaths due to health care demand exceeding health care capacity. However, fears of hospital overcrowding persist [26].

## Results

As of May 25, a total of 129,148 cases and 7,660 deaths due to CoVID-19 have been reported by the Ministry of Health, Peru. Among men, reported cases were mostly observed among individuals aged 30–39 years (23.1%), followed by those aged 40–49 years (20.8%), and those aged 20–29 years (18.3%). In contrast, most deaths were reported among those aged 50 years and above, especially among men aged 60–69 (29.3%) followed by those aged 70–79 (23.2%), aged 50–59 years (20.1%), and aged 80 years and above (14.3%). (Table 1, Figure 1A, 1B). Data show a similar pattern for women. The majority of reported cases occur in females aged 20–69 years, and the majority of reported deaths occur among women aged 50 years or more. More specifically, most reported cases occur among women aged 30–39 (22.6%), followed by women aged 40–49 (19.7%), and 50–59 year olds (16.0%). In contrast, most deaths are reported among those aged 60–69 (30.2%), followed by women aged 70–79 (25.9%), and lastly, women aged 80 years and above (18.4%). Regarding CoVID-19 mortality per 100,000 population, seniors (individuals > 70 years of age) were the most affected age group; mortality burden per 100,000 is 279.2 among men aged 80 years and above, and 207.6 among men aged 70–79 years. For women of 80 years of age or more mortality is 108.8 and 85.1 for women aged 70–79 years (Table 1, Figure 1F).

**Table 1.** Distribution of the cases by sex and age groups, as of May 25, 2020.

| Age group | Men |  |  |  | Women |  |  |  |
| --- | --- | --- | --- | --- | --- | --- | --- | --- |
|  | Cases<br>(%) | Deaths<br>(%) | cCFR<br>(%) | Mortality<br>per 100,000<br>population | Cases<br>(%) | Deaths<br>(%) | cCFR(%)<br>) | Mortality<br>per 100,000<br>population |
| All | 78264<br>(100) | 5508<br>(100) | 7.0% | 34.0 | 50884<br>(100) | 2152<br>(100) | 4.2 | 13.1 |
| 0-9 | 1416<br>(1.8) | 10<br>(0.2) | 0.7 | 0.4 | 1362<br>(2.7) | 11<br>(0.5) | 0.8 | 0.4 |
| 10-19 | 2475<br>(3.2) | 8<br>(0.1) | 0.3 | 0.3 | 2128<br>(4.2) | 6<br>(0.3) | 0.3 | 0.2 |
| 20-29 | 14306<br>(18.3) | 32<br>(0.6) | 0.2 | 1.2 | 8707<br>(17.1) | 19<br>(0.9) | 0.2 | 0.7 |
| 30-39 | 18052<br>(23.1) | 169<br>(3.1) | 0.9 | 6.6 | 11487<br>(22.6) | 49<br>(2.3) | 0.4 | 2.0 |
| 40-49 | 16258<br>(20.8) | 499<br>(9.1) | 3.1 | 23.7 | 10005<br>(19.7) | 140<br>(6.5) | 1.4 | 6.7 |
| 50-59 | 13274<br>(17.0) | 1107<br>(20.1) | 8.3 | 67.7 | 8124<br>(16.0) | 323<br>(15.0) | 4.0 | 19.7 |
| 60-69 | 7034<br>(9.0) | 1615<br>(29.3) | 23.0 | 150.4 | 5023<br>(9.9) | 649<br>(30.2) | 12.9 | 56.6 |
| 70-79 | 3620<br>(4.6) | 1279<br>(23.2) | 35.3 | 207.6 | 2488<br>(4.9) | 558<br>(25.9) | 22.4 | 85.1 |
| 80 - | 1769<br>(2.3) | 789<br>(14.3) | 44.6 | 279.2 | 1536<br>(3.0) | 397<br>(18.4) | 25.8 | 108.8 |

**Figure 1.**
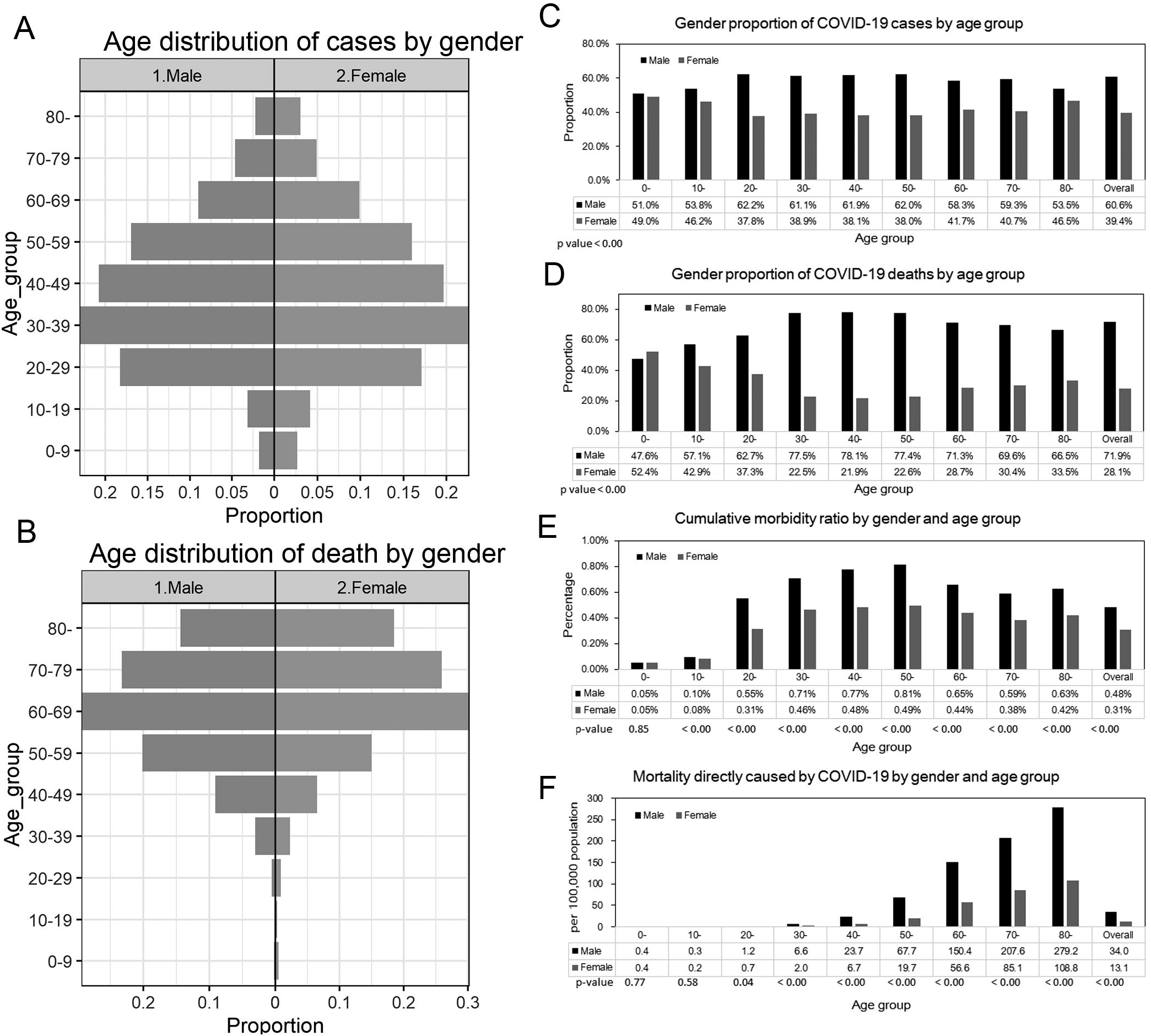
Epidemiological characterization of COVID-19 in Peru, as of May 25, 2020 (A) Age distribution of reported cases by gender, (B) Age distribution of reported deaths by gender. (C) Gender proportion of COVID-19 cases by age group, (D) Gender proportion of COVID-19 deaths by age group, (E) Cumulative morbidity ratio by gender and age group, (F) Mortality directly caused by COVID-19 by gender and age group

The gender proportions of reported cases by age groups are presented in Figure 1C and Figure 1D. The proportion of cases among men is higher than 50% across all age groups (χ^2^ test, p-value<0.001). Similarly, the proportion of male deaths is also higher than 50% except for those aged 10–19 years (χ^2^ test, p-value<0.001). Cumulative morbidity ratio by gender and age group is presented in Figure 1E, indicating that cumulative morbidity ratio among men is higher than women across all age groups (proportion test, p-value < 0.001) except for individuals aged 0–9 years (proportion test, p-value = 0.85). Figure 1F illustrates the mortality per 100,000 population directly caused by CoVID-19 by gender and age group. Mortality is higher than among females aged 20 years and above (proportion test, p-value <0.05), and it is not significantly different among those aged 0–19 years.

Figure 2 shows the cumulative cases and deaths of CoVID-19 by age group for males and females (A through J) over time. The figure suggests cumulative deaths increases after an increase in cumulative cases. The growth curve for overall cumulative cases (all age groups) for men and women appears to increase exponentially until around day 60 (April 29^th^, 2020), while exponential growth in cumulative deaths overall (all age groups) for men and women appears to occur until around day 70 (May 9^th^, 2020).

**Figure 2:**
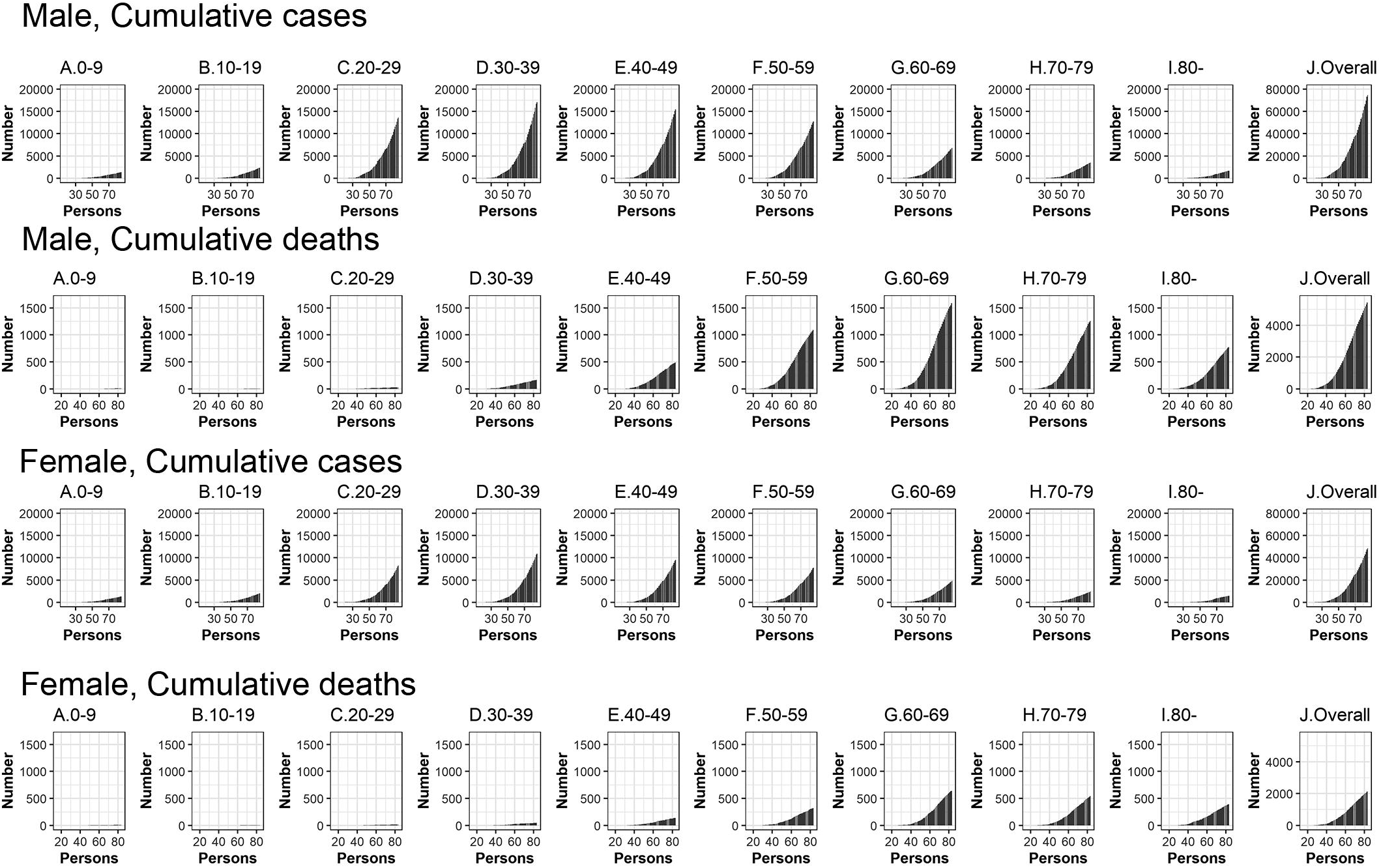
Temporal distribution of cases and deaths by age group due to COVID-19, March-May 2020, Peru. Top: Male, cumulative cases, Second top: Male, cumulative cases, Second bottom: Female, cumulative cases, Bottom: Female cumulative deaths (A) aged 0–9, (B) aged 10–19, (C) aged 20–29, (D) aged 30–39, (E) aged 40–49, (F) aged 50–59, (G) aged 60–69, (H) aged 70–79,(I) aged 80- and (J) Overall (all age groups). Day 1 corresponds to March 1st in 2020.

Figure 3 illustrates observed and model based posterior estimates of the crude CFR by age group (A-J) and time-delay adjusted CFR by age group (K-T) for men and women. Black dots show crude case fatality ratios, and light and dark indicate 95% and 50% credible intervals (CrI) for posterior estimates, respectively.

**Figure 3:**
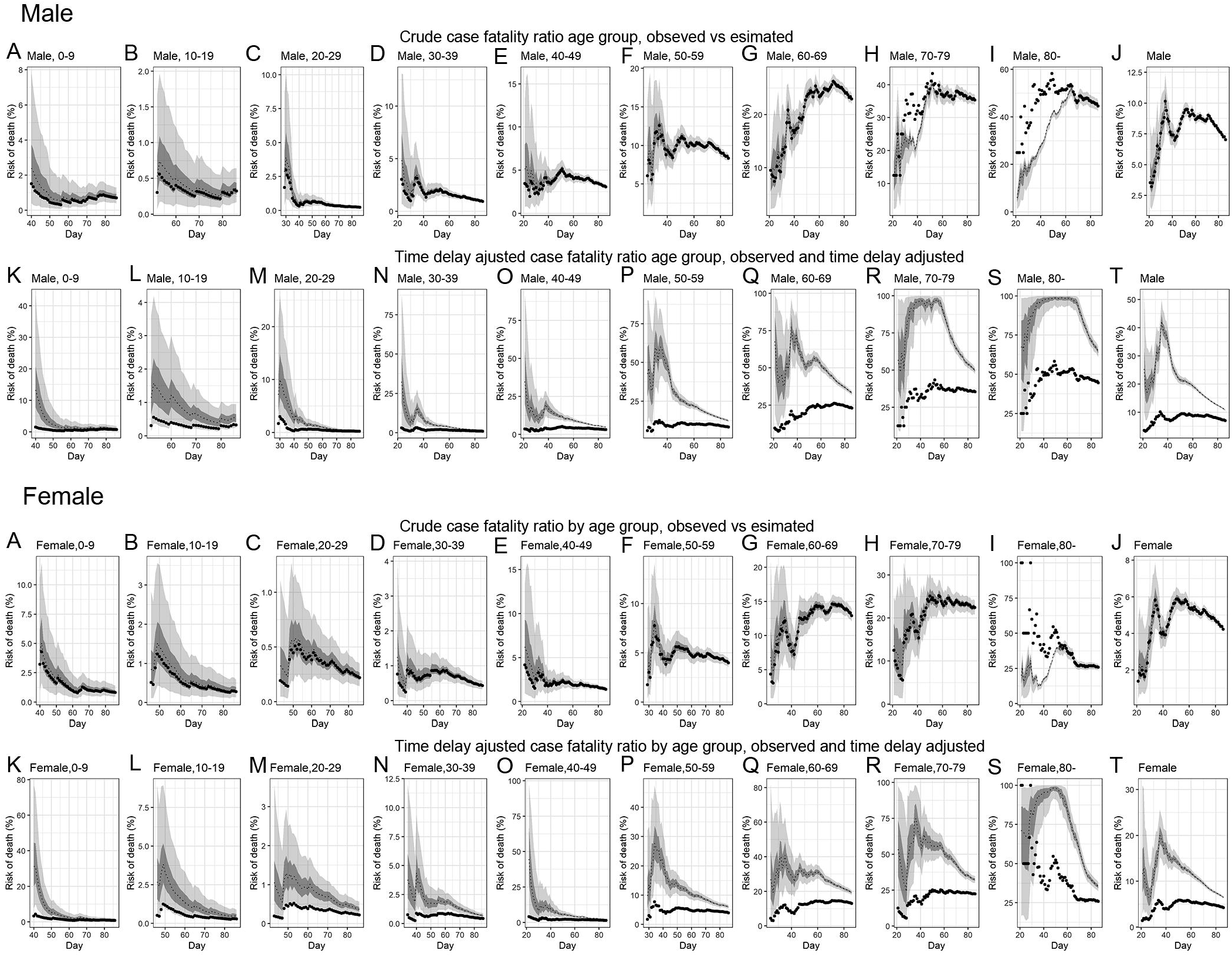
Temporal variation of male and female risk of death by age group caused by COVID-19, March-May 2020, Peru. Upper two rows; Male risk of deaths, Lower two rows; Female risk of deaths Observed and posterior estimated of crude case fatality ratio of (A) aged 0–39, (B) aged 40–49, (C) aged 50–59, (D) aged 60–69, (E) aged 70–79, (F) aged 80-, (G) all age groups and time-delay adjusted case fatality ratio of (H) aged 0–39, (I) aged 40–49, (J) aged 50–59, (K) aged 60–69, (L) aged 70–79, (M) aged 80-, (N) Overall (all age groups). Day 1 corresponds to March 1st in 2020. Black dots show crude case fatality ratio, and light and dark indicates 95% and 50% credible intervals for posterior estimates, respectively.

Overall, our model based crude CFR fitted the observed data well, except for individuals aged 80 years and above, probably influenced by low reporting rate/ascertainment bias of cases at an early stage. Crude CFR for most of age groups increased at the early stage of the epidemic, peaked amidst the outbreak ∼day 34 (April 3^rd^, 2020) and followed a decreasing trend turning into an almost flat curve.

Overall, our model-based posterior estimates for the time-delay adjusted CFR are substantially higher than the crude observed CFR. These estimates fluctuated at the early stage of the epidemic and then followed a decreasing trend.

The most recent estimates, as of May 25, 2020, of the time-delay adjusted CFR for men and women are 10.8% (95%CrI: 10.5–11.1%) and 6.5% (95%CrI: 6.2–6.8%), respectively, while overall national estimate is 9.1% (95%CrI: 8.9–9.3%) (Figure 4 and Table 2). Among men, senior citizens appear to be severely affected; the adjusted CFR is 33.1% (95%CrI: 31.7–34.6%) for men aged 60–69 years, 49.4% (95%CrI: 47.3–51.6%) for those aged 70–79 years, and 64.3% (95%CrI: 60.9–67.8%) for those 80 years old and above. We observe a similar pattern for women. The adjusted CFR is 19.2% (95%CrI: 17.9–20.6%) for women aged 60–69 years, 32.2% (95%CrI: 29.9–34.7%) for those aged 70–79 years, and 35.1% (95%CrI: 32.1–38.1%) for women aged 80 years old or more.

**Figure 4.**
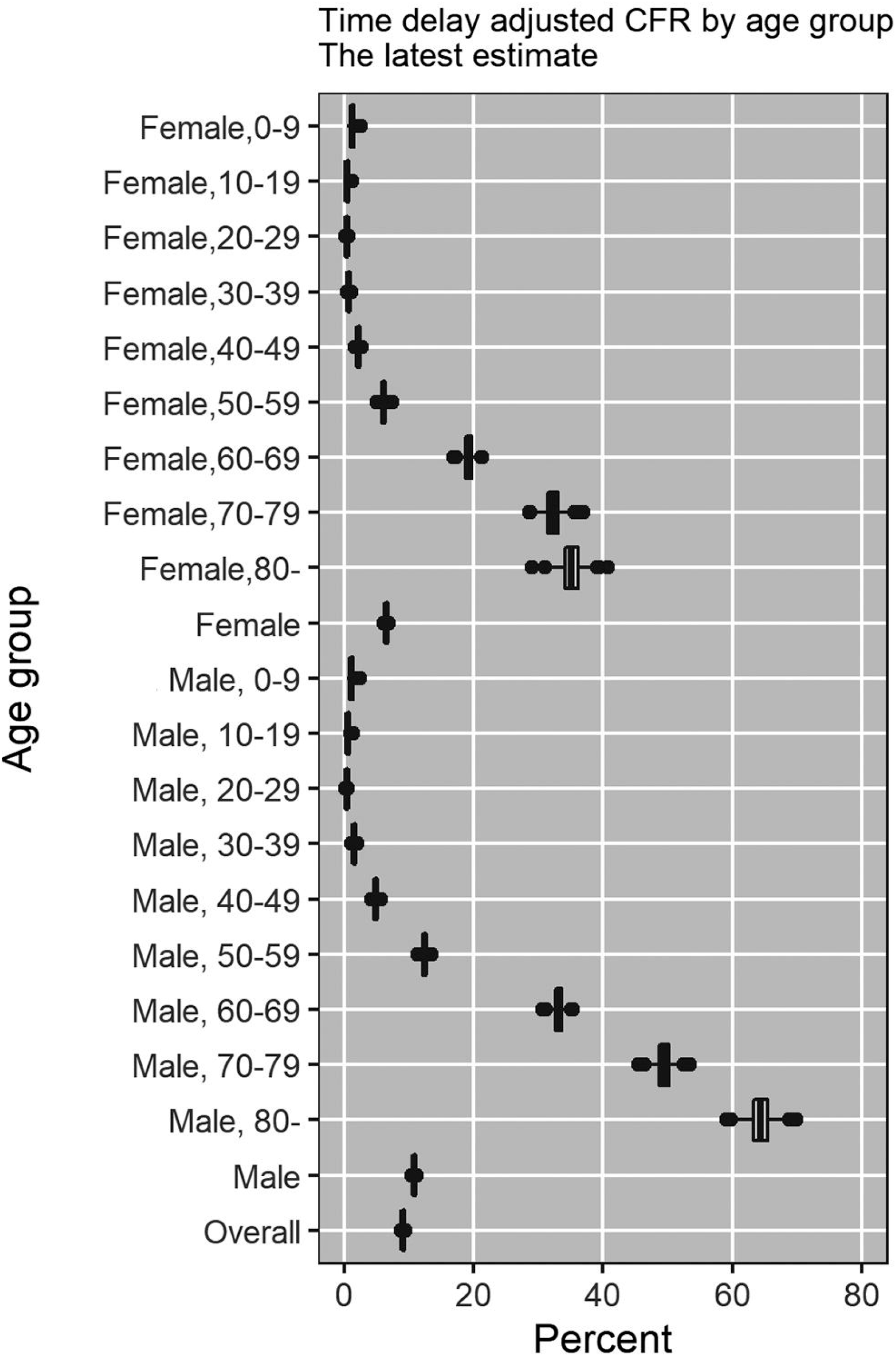
Latest estimates of time-delay adjusted risk of death caused by COVID-19 by age group and gender, March-May 2020, Peru. Distribution of time-delay adjusted risk of death from the latest estimates (May 25, 2020) is presented. Top to bottom: female aged 0–9, female aged 10–19, female aged 20–29, female aged 30–39, female aged 40–49, female aged 50–59, female aged 60–69, female aged 70–79, female aged 80 and over, female overall

**Table 2.** Summary results of time-delay adjusted case fatality ratio of COVID-19 in each age group in Peru, 2020 as of May 25, 2020.

| Age group | Gender | Latest estimate | Range of median estimates | Crude case fatality rate |
| --- | --- | --- | --- | --- |
| Overall |  | 9.1% (95%CrI<sup>a</sup>: 8.9-9.3%) | 9.1-32.0% | 5.9% (95%CI<sup>b</sup>: 5.8-6.1%)<br>7660/129148<sup>c</sup> |
| Male |  | 10.8% (95%CrI: 10.5-11.1%) | 10.8-42.3% | 7.0% (95%CI: 6.9-7.2%)<br>5508/78264 |
| Female |  | 6.5% (95%CrI: 6.2-6.8%) | 6.4-20.0% | 4.2% (95%CI: 4.1-4.4%)<br>2152/50884 |
| 0-9 | Male | 1.1% (95%CrI: 0.5-1.8%) | 1.0-13.3% | 0.7% (95%CI: 0.3-1.3%)<br>10/1416 |
|  | Female | 1.2% (95%CrI: 0.7-2.0%) | 1.2-31.4% | 0.8% (95%CI: 0.4-1.4%)<br>11/1362 |
| 10-19 | Male | 0.5% (95%CrI: 0.3-1.0%) | 0.4-1.6% | 0.3% (95%CI: 0.1-0.6%)<br>8/2475 |
|  | Female | 0.5% (95%CrI: 0.2-0.9%) | 0.4-3.8% | 0.3% (95%CI: 0.1-0.6%)<br>6/2128 |
| 20-29 | Male | 0.4% (95%CrI: 0.2-0.5%) | 0.4-10.0% | 0.2% (95%CI: 0.2-0.3%)<br>32/14306 |
|  | Female | 0.4% (95%CrI: 0.2-0.6%) | 0.4-1.3% | 0.2% (95%CI: 0.1-0.3%)<br>19/8707 |
| 30-39 | Male | 1.5% (95%CrI: 1.3-1.7%) | 1.5-32.3% | 0.9% (95%CI: 0.8-1.1%)<br>169/18052 |
|  | Female | 0.7% (95%CrI: 0.5-0.9%) | 0.9-4.4% | 0.4% (95%CI: 0.3-0.6%)<br>49/11487 |
| 40-49 | Male | 4.8% (95%CrI: 4.4-5.2%) | 4.8-34.7% | 3.1% (95%CI: 2.8-3.3%)<br>499/16258 |
|  | Female | 2.2% (95%CrI: 1.8-2.5%) | 2.2-44.6% | 1.4% (95%CI: 1.2-1.6%)<br>140/10005 |
| 50-59 | Male | 12.4% (95%CrI: 11.7-13.1%) | 12.4-60.0% | 8.3% (95%CI: 7.9-8.8%)<br>1107/13274 |
|  | Female | 6.0% (95%CrI: 5.4-6.7%) | 7.5-27.7% | 4.0% (95%CI: 3.6-4.4%)<br>323/8124 |
| 60-69 | Male | 33.1% (95%CrI: 31.7-34.6%) | 33.1-77.8% | 23.0% (95%CI: 22.0-24.0%)<br>1615/7034 |
|  | Female | 19.2% (95%CrI: 17.9-20.6%) | 19.2-40.9% | 12.9% (95%CI: 12.0-13.9%)<br>649/5023 |
| 70-79 | Male | 49.4% (95%CrI: 47.3-51.6%) | 48.7-97.8% | 35.3% (95%CI: 33.8-36.9%)<br>1279/3620 |
|  | Female | 32.2% (95%CrI: 29.9-34.7%) | 24.1-74.9% | 22.4% (95%CI: 20.8-24.1%)<br>558/2488 |
| 80- | Male | 64.3% (95%CrI: 60.9-67.8%) | 64.3-98.9% | 44.6% (95%CI: 42.3-47.0%)<br>789/1769 |
|  | Female | 35.1% (95%CrI: 32.1-38.1%) | 35.1-98.3% | 25.8% (95%CI: 23.7-28.1%)<br>397/1536 |

<sup>a</sup> 95%CrI: 95% credibility intervals (CrI), <sup>b</sup> 95%CI: 95% confidence interval, <sup>c</sup> Cumulative cases over cumulative deaths

## Discussion

This study estimates the time-delay adjusted CFR by age group for the ongoing CoVID-19 epidemic in Peru. The crude CFR varies across countries due to differences in testing and timing of tests [27]. The results show that the CoVID-19 epidemic in Peru disproportionately impacts senior individuals, especially those who are 70 years of age or older, consistent with CFR estimates obtained from recent studies conducted in China [28, 29], Chile [30], and Italy [31, 32]. This pattern, suggests that an aging population could aggravate the fatality impact of CoVID-19, similar to influenza and respiratory syncytial virus [30], as was probably the case in Italy [31, 32]. While the population in Lain America, including Peru, is aging at a rapid rate, still a relatively small percentage of the population in the region are older than 65 years of age [33]. So in fact the age structure in the region could favor a lower overall CFR than would be expected otherwise with a relatively older population, as in other regions.

Our estimate of adjusted CFR among men (10.8% (95%CrI: 10.5–11.1%)) is 1.7-fold higher than the estimated adjusted CFR for women (6.5% (95%CrI: 6.2–6.8%)), consistent with the estimates given in ref [34]. Men aged 80 years or older have an estimated adjusted CFR as high as 64.3% (95%CrI: 60.9–67.8%), 58-fold higher than our estimates for men aged 0–9, and 1.3-fold higher than our estimates for men aged 70–79. Similarly, the adjusted CFR estimates for women of aged 80 years or older are as high as 35.1% (95%CrI: 32.1–38.1%), 29-fold higher than the estimates obtained for female aged 0–9 and 1.1-fold higher than the estimates obtained for female aged 70–79, consistent with recent findings in Chile [30]. In comparison, a study conducted in China, reported much lower estimates of CFR for individuals >80 years of age (13.4%) [29].

An upward trend in the crude CFR for overall population suggests the transmission may be spreading to more vulnerable populations. The majority of social distancing measures in Peru were implemented between March 11-March 18, 2020. However, since 72.4% of the economically active population works in informal jobs, which are concentrated in the poorest areas of the country, compliance with government mitigation strategies can be challenging despite the government’s efforts to support the population [35]. Another factor possibly contributing to the upward trend in crude CFR may be an increase in unreported cases due to saturated testing capacity [27]. However, since Peru’s testing capacity has substantially increased since the beginning of the outbreak, going from >0.01 test per 1000 population to 0.09 per 1000 in May 22 [15], and the positivity rate is estimated at ∼8.6% as of March 30, 2020, this seems an unlikely cause. Furthermore, the results show an increasing trend in crude CFR around day 45 (May 14^th^, 2020), probably reflecting the exponential increase of cumulative cases around day 40 (May 9^th^, 2020).

The downward trend in the adjusted CFR at the early stage may indicate the existence of a reporting delay and the shift of the outbreak to a less vulnerable segment of the population. In particular, the observed differences in estimates between the crude CFR and adjusted CFR can be attributed to the time-delay that is assumed fixed during the course of the epidemic.

The relatively small proportion of males (53.5%) among CoVID-19 cases in the individuals aged 80 years and above can be attributed to the relatively small male population size for that age group; with men comprising only 1.7% of the population > 80 years of age in Peru, consistent with estimates for Chile [30]. As higher mortality among male has been reported in China and the U.S. [34], additional data on deaths stratified by gender provides the opportunity to examine the CFR by gender and age.

Our study has at least two limitations. First, our estimates are probably overestimated, due to the effect of under reporting rates and ascertainment rates, as has been underscored in other studies [22, 24, 36]. But a recently enhanced testing capacity in Peru is expected to mitigate these effects, and an ongoing mass serological study will provide data to generate more accurate estimates of the death risk. Second, adjusted CFR, especially among seniors, has displayed fluctuations, highlighting the importance of focusing on sub-group analyses. Additional information such as line lists that include related risks including information on underlying diseases may help to identify subgroups with elevated risks.

## Conclusions

The COVID-19 epidemic is imposing a large death toll in Peru. Senior individuals, especially those who are older than 70 years of age, are being disproportionately affected by the COVID-19 pandemic, particularly elderly men. CFR was as high as 64.3% (95%CrI: 60.9–67.8%) for men aged 80 older, 58-fold higher than our estimates for men aged 0–9. The overall adjusted CFR is larger than in other countries, which is worrying, particularly because healthcare demand has not yet exceeded capacity, but probably will do in the coming weeks. The relatively younger age structure in Latin America may help ameliorate the overall CFR than would otherwise be expected with an older age structure in the population.

## Data Availability

The data that support the findings of this study are available from the Ministry of Health in Peru. Data was anonymized prior to the authors gaining access. Restrictions apply to the availability of these data, which were used under license for this study. Data are available from the authors with the permission of the Ministry of Health in Peru.

## Materials and methods

### Data

We obtained daily cumulative numbers of reported laboratory confirmed CoVID-19 cases and deaths stratified by age group and gender through May 25, 2020. Different age groups had different starting times, which correspond to the day when death was reported. Confirmed CoVID-19 cases were retrieved from three surveillance systems: a) national surveillance system (confirmed and suspected cases based on a case definition), b) Netlab system (molecular test) and c) SICOVID system (rapid serological test). COVID-19 deaths were obtained from two surveillance systems: a) national surveillance system (confirmed and suspected deaths based on a case definition) and b) Vital statistics system (National System of mortality-SINADEF-which is an online system that keeps track of death certificates).

Population size by age, group, and gender in 2020 were retrieved from the Ministry of Health in Peru [37]

### Statistical analysis

The crude CFR is defined as the number of cumulative deaths over the number of cumulative cases. For the estimation of CFR in real time, we employed the delay from hospitalization to death, h_*s*_, which is assumed to be given by h_*s*_ = H(s) – H(s–1) for s>0 where H(s) is a cumulative density function of the delay from hospitalization to death and follows a gamma distribution with mean 10.1 days and SD 5.4 days, as given in ref, Mizumoto and Chowell [21]. Let π_*a,ti*_ be the time-delay adjusted case fatality ratio on reported day t_*i*_ in area *a*, the likelihood function of the estimate π*_a,t_i__* is given by equation:

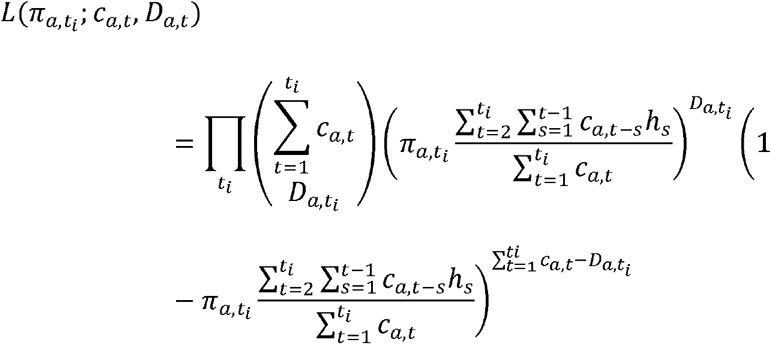

where c_*a,t*_ represents the number of new cases with reported day t in area *a*, and *D_a,t_i__* is the cumulative number of deaths until reported day t_*i*_ in area *a* [38, 39]. Among the cumulative cases with reported day *t* in area *a*,*D_a,t_i__* have died and the remainder have survived the infection. The contribution of those who have died with biased death risk is shown in the middle parenthetical term and the contribution of survivors is presented in the right parenthetical term. We assume that *D_a,t_i__*is the result of the binomial sampling process with probability π*_a,t_i__*

We used a Monte Carlo Markov Chain (MCMC) method in a Bayesian framework to estimate model parameters. We evaluated the convergence of MCMC chains using the potential scale reduction statistic [40, 41]. Estimates and 95% credibility intervals for these estimates are based on the posterior probability distribution of each parameter and samples drawn from the posterior distributions. All statistical analyses were conducted in R version 3.6.1 (R Foundation for Statistical Computing, Vienna, Austria) using the ‘rstan’ package.

## Acknowledgments

This work was supported by the Japan Society for the Promotion of Science (JSPS) KAKENHI [grant Number 20H03940]; the Leading Initiative for Excellent Young Researchers from the Ministry of Education, Culture, Sport, Science & Technology of Japan to [KM]; National Science Foundation NSF [grant 1414374] as part of the joint NSF-National Institutes of Health NIH-United States Department of Agriculture USDA Ecology and Evolution of Infectious Diseases program; UK Biotechnology and Biological Sciences Research Council [grant BB/M008894/1] to [GC]; and the ANID Millennium Science Initiative/ Millennium Initiative for Collaborative Research on Bacterial Resistance, MICROB-R, [NCN17_081] to [EU].

## Author Contributions

GC and KM conceived the early study idea. KM implemented statistical analysis. AT and KM wrote the first full draft. CM performed data acquisition. All authors contributed to the revision of the manuscript. AU advised the study, and revised the manuscript. GC advised on and helped shape the research. All authors contributed to the interpretation of the results and edited and commented on several earlier versions of the manuscript. All authors read and approved the final manuscript.

## Conflict of interest

All authors report no conflicts of interest.

## Notes

### Competing Interest Statement

The authors have declared no competing interest.

